# Machine learning based on event-related oscillations of working memory differentiates between preclinical Alzheimer’s disease and neurotypical aging

**DOI:** 10.1101/2023.10.11.23296890

**Authors:** Ke Liao, Laura E. Martin, Sodiq Fakorede, William M. Brooks, Jeffrey M. Burns, Hannes Devos

## Abstract

There is increasing evidence of the usefulness of electroencephalography (EEG) as an early neurophysiological marker of preclinical AD. Our objective was to apply machine learning approaches on event-related oscillations to discriminate preclinical AD from neurotypical controls. Twenty-two preclinical AD participants who were cognitively normal with elevated amyloid and 21 cognitively normal with no elevated amyloid controls completed n-back working memory tasks (n= 0, 1, 2). EEG signals were recorded through a high-density sensor net. The event-related spectral changes were extracted using the discrete wavelet transform in the delta, theta, alpha, and beta bands. The support vector machine (SVM) machine learning method was employed to classify participants, and classification performance was assessed using the Area Under the Curve (AUC) metric. The relative power of the beta and delta bands outperformed other frequency bands with higher AUC values. The 2-back task obtained higher AUC values than the 0 and 1-back tasks. The highest AUC values were from the 2-back task beta band (AUC = 0.86) and delta bands (AUC = 0.85) nontarget data. This study demonstrates the promise of using machine learning on EEG event-related oscillations from working memory tasks to detect preclinical AD.

## 1. Introduction

Alzheimer’s disease (AD) is one of the most common neurodegenerative diseases and the leading cause of dementia worldwide. About 50 million people were living with dementia and related disorders in 2018, and the prevalence is expected to increase to more than 150 million by the year 2050, according to the World Alzheimer Report (Patterson, 2018). This projected prevalence puts a tremendous burden on the health, long-term care, and hospice services for people with AD and dementia (Alzheimer’s Association, 2023).

Electroencephalography (EEG) is an electrophysiological technique for recording brain electrical activity, which is a summation of the excitatory and inhibitory postsynaptic potentials of relatively large groups of neurons firing synchronously (Britton et al., 2016). EEG has been adopted for studying the neural dynamics across all stages of AD (Horvath et al., 2018; Maestú et al., 2019; Ouchani et al., 2021; Perez-Valero et al., 2021; Smailovic & Jelic, 2019). The high temporal sensitivity, non-invasiveness, cost-effectiveness, and accessibility make EEG suitable for evaluating dynamic brain functioning and screening for early AD. These features of EEG may complement existing biomarkers such as tracing amyloid beta (Aβ) and tau in the blood, cerebrospinal fluid (CSF), or brain; assessing cortical hypometabolism on fluorodeoxyglucose positron emission tomography (FDG-PET); and evaluating brain atrophy on structural magnetic resonance imaging (MRI) (Jack et al., 2013; Jack et al., 2010).

Most EEG studies have focused on detecting changes in neural dynamics in people with diagnosed AD or mild cognitive impairment (MCI) (Ishii et al., 2017; Poza et al., 2014; Rossini et al., 2007). Individuals in these stages of AD already have marked cognitive impairments that may or may not affect daily life activities. However, few have investigated the preclinical phase of AD, which is characterized by early pathological changes related to AD in the presence of normal cognition (Gouw et al., 2017; Nakamura et al., 2018; Stomrud et al., 2010). Specifically, Stomrud et al. (2010) reported that total tau and phosphorylated tau (P-tau), and the combined P-tau/Aβ42 ratio measured in CSF correlated with relative EEG theta power, and slowing of cognitive speed correlated with increased relative theta power and high P-tau/Aβ42 ratio in cognitively normal older adults. Gouw et al. (2017) found that in participants with subjective cognitive decline, higher delta and theta power, and lower alpha power and peak frequency were associated with clinical progression over at least one year. Nakamura et al. (2018) demonstrated that in older adults with MCI and controls, increased Aβ levels were associated with higher alpha power in the medial frontal areas. In addition, increased delta power in the same region was associated with disease progression, entorhinal atrophy, and regional decrease in glucose metabolism.

The EEG power spectrum change is a promising neurophysiological biomarker to characterize the neural activity alterations associated with AD progression (Dauwels et al., 2011; Moretti, 2016; Stam et al., 2005). Spectral analysis can be applied to either resting-state EEG collected while participants are not doing any purposeful activity (Cassani et al., 2018; Moretti et al., 2004; Trinh et al., 2021) or during tasked-based EEG while participants perform a cognitive task (Chapman et al., 2011; Karrasch et al., 2006; Lai et al., 2010; Missonnier et al., 2007). The spectral changes under resting-state condition have been used successfully as features in studies on preclinical AD (Gouw et al., 2017; Nakamura et al., 2018; Stomrud et al., 2010). Spectral analysis for task-based studies have examined the event-related desynchronization/synchronization (ERD/ERS) (Babiloni et al., 2020; Fraga et al., 2018) and event-related oscillations (ERO) (Başar et al., 2013; Yener & Başar, 2013). Participants with MCI exhibited lower delta band oscillatory responses than neurotypical controls in oddball paradigms (Kurt et al., 2014; Yener et al., 2013). Participants with amnestic MCI demonstrated significantly lower theta power in a Sternberg recognition task than neurotypical controls (Cummins et al., 2008). Likewise, induced theta activity was significantly lower in progressive MCI than controls and stable MCI using n-back working memory tasks (Deiber et al., 2009). However, whether these task-related power spectrum rhythms also demonstrate in older adults with preclinical AD is unclear.

Machine learning offers a systematic approach to developing sophisticated, automatic, and objective classification frameworks for analyzing and recognizing complex and subtle patterns of high-dimensional data. Machine learning has been used in many MCI and AD studies (Mirzaei & Adeli, 2022; Pellegrini et al., 2018; Rathore et al., 2017; Tanveer et al., 2020) since it holds promise for improving the sensitivity and/or specificity of disease diagnosis (Rathore et al., 2017; Sajda, 2006). Among them, Petrosian et al. (2001) trained recurrent neural networks (RNN) on resting-state EEG and obtained an accuracy under 90% in classifying participants with AD and controls. Fiscon et al. (2018) used decision trees classifiers on resting-state EEG and achieved 83%, 92%, and 79% accuracy in categorizing AD vs. controls, MCI vs. controls, and MCI vs. AD groups, respectively. You et al. (2020) proposed a cascade neural network on resting-state EEG with gait data and reached an accuracy rate of 91% in the three-way classification of controls, MCI, and AD. Khatun et al. (2019) used support vector machine (SVM) classifiers to predict MCI vs controls with 88% accuracy on auditory evoked potentials. However, no studies have used machine learning based on EEG oscillatory changes to classify participants with preclinical AD and controls.

In this study, we proposed to use event-related oscillations from working memory tasks, instead of resting-state EEG, to differentiate preclinical AD from neurotypical aging since previous studies have shown that memory activations can perform better than resting-state EEG in revealing spectral differences between participants with MCI and controls (van der Hiele et al., 2007). We hypothesized that machine learning approaches would detect early changes in spectral frequencies associated with AD and categorize participants with preclinical AD and controls with high accuracy.

## 2. Materials and Methods

### 2.1 Participants

The study cohort consisted of two groups of cognitively normal participants: 21 controls with no elevated Aβ and 22 preclinical AD adults with elevated Aβ. All participants were recruited exclusively from the University of Kansas Alzheimer’s Disease Research Center during the period of May 30, 2018 to September 17, 2021. Participants were ineligible if they met any of the following criteria based on their medical records: (1) currently using steroids, benzodiazepines, or neuroleptics; (2) had a history of substance abuse; (3) had a previous neurological disorder; or (4) had any contraindications to PET or EEG. The inclusion criteria consisted of the following: (1) being 65 years of age or older; (2) having a comprehensive understanding of all instructions given in English; (3) providing informed consent; and (4) having previously undergone an amyloid PET scan of the brain. The cerebral amyloid burden was determined using PET images acquired from a GE Discovery ST-16 PET/CT scanner subsequent to the administration of intravenous Florbetapir 18F-AV45 (370MBq), following a previously published protocol (Vidoni et al., 2021). Aβ status was determined by three experienced raters who independently interpreted all PET images without access to any clinical information, employing a method described in previous studies (Harn et al., 2017). The raters employed a combination of visual and quantitative data to establish whether a participant was classified as non-elevated or elevated (Clark et al., 2012; Joshi et al., 2012). The final status was determined based on the majority decision among the three raters. The median duration between the PET scan and EEG assessment was 1047 (823 - 1578) days. The participants’ demographic information is shown in Table 1. The two groups did not differ with respect to age (p = .60), but there was a significant difference (p < .01) in the Montreal Cognitive Assessment (MoCA) scores (Nasreddine et al., 2005) with the MoCA scores for the control group (mean = 28.10) being higher than the preclinical AD group (mean = 26.05).

**Table 1.** Demographic information of study participants. Continuous variables were expressed as mean with standard deviation. The bold values were significant at p < .01 under the two-sample unequal variance t-test.

| Group | Number | Sex (male: female) | Age | MoCA |
| --- | --- | --- | --- | --- |
| Controls | 21 | 8:13 | 74.38(5.93) | **28.10(1.89)** |
| Preclinical AD | 22 | 6:16 | 73.52(4.52) | **26.05(2.79)** |

### 2.2 Data acquisition and processing

The n-back working memory task (Fraga et al., 2018; Missonnier et al., 2006) was used to evoke event-related potentials. The participants were visually presented with a series of letters and instructed to respond in target condition trials when the current letter was ‘x’ in the 0-back task or was the same as the one presented in n (n = 1, 2) trials previously. In nontarget condition trials, participants were instructed to withhold responses (Devos et al., 2022; Devos et al., 2023).

The EEG signals were collected with a 256-channel high-density sensor net (Magstim EGI, Eugene, OR) at a sampling frequency of 1000 Hz and online referenced to the Cz channel. In offline processing, a digital filter of 0.5 to 30 Hz was applied, and the linked mastoid re-referencing was used. Non-neural activities were identified and removed using independent component analysis in EEGLAB (Delorme & Makeig, 2004). The clean data had 184 EEG channels with epoch length from -100ms to 1000ms in regard to stimulus onset. Only the trials with correct behavioral responses were included in the analysis. For more data cleaning and processing details, please see the original publications (Devos et al., 2022; Devos et al., 2023).

### 2.3 Spectral analysis

The individual averaged data were used for spectral analysis, and the wavelet-based relative power was obtained on each EEG channel. Wavelet transform is a time-frequency representation of the signal, which decomposes signals into different subbands (levels) with signal approximation and details. Compared to other spectral analyses like fast Fourier transform, wavelet transform can keep both the temporal and frequency information of the signal. As EEG is a non-stationary signal with changing properties over time, wavelet transform can catch the signal’s transient features due to the scalability of its analyzing window (Strang & Nguyen, 1996). It enriches the analysis with more details than conventional time-frequency algorithms, such as short-time Fourier transform. Therefore, wavelet transform is a well-established signal representation and feature extraction technique for EEG processing (Faust et al., 2015; Frantzidis et al., 2014; Murugappan et al., 2010; Petrosian et al., 2001).

This study employed a multi-resolution decomposition method based on discrete wavelet transform (DWT) in power spectral analysis. In practice, DWT uses a low-pass filter *h*(*n*) and a high-pass filter *g*(*n*) to decompose the signal into its low-pass part (approximation) with its high-pass part (detail). In multi-level decomposition, DWT recursively decomposes approximation coefficients with the same pair of low and high-pass filters to the desired levels. The features extracted from the approximation and detailed coefficients at various levels can reveal the characteristics of the analyzed time series at different frequency bands. Figure 1 shows a schematic implementation of the DWT multi-resolution analysis using low-pass and high-pass filters.

**Figure 1.**
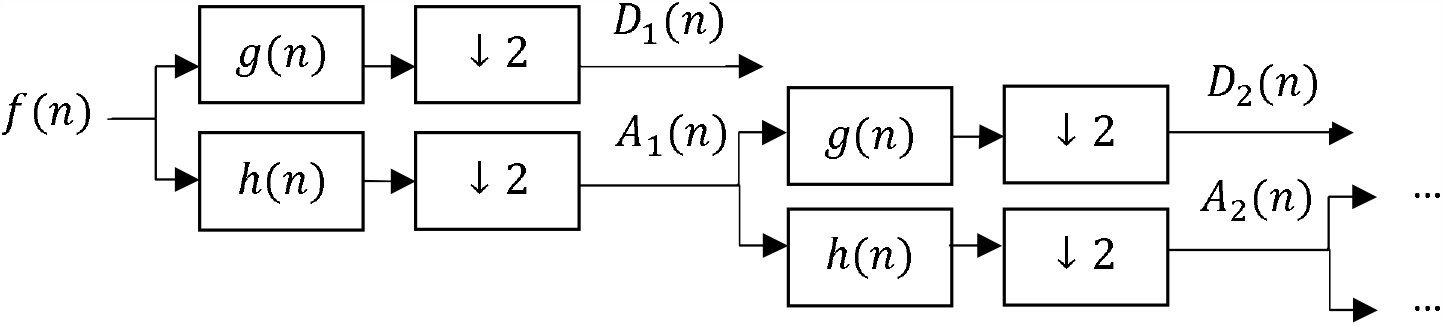
Discrete wavelet transform implementation using low-pass and high-pass filters.

In Figure 1, only two levels of DWT decomposition were shown. The operation ↓2 denoted a decimation of the signal by a factor of two.

Let *D*_*i*_(*n*) and *A*_*i*_ (*n*) denote the detailed component and the approximate component on the decomposition level *i*, and the maximum level is *M*. The original EEG signal *f*(*n*) could be represented as the sum of wavelet-based subbands in a multi-resolution analysis as

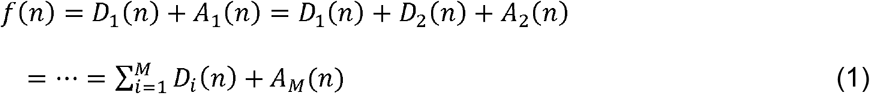

Where *A*_*i*_(*n*) *= A*_*i*+l_(*n*) *+ D*_*i*+l_(*n*) in two consecutive levels *i* and *i* + 1.

A seven-level discrete wavelet transform decomposition (*M* = 7) was applied to the EEG signals. Only the DWT subbands obtained from the fifth to seventh levels were used in the analysis.

Table 2 lists the decomposed signals A7, D7, D6, D5, which roughly corresponded to the EEG signal frequency bands of delta, theta, alpha, and beta, respectively. Accordingly, equation (1) could be calculated as follows:

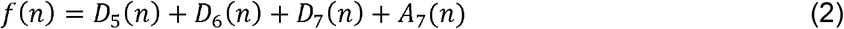

For each of the frequency bands mentioned above, the DWT relative spectral power *RP*_*i*_ (*i* = 4, 5, 6, 7) was obtained as follows (Rosso et al., 2006; Wan et al., 2006; Wang et al., 2016):

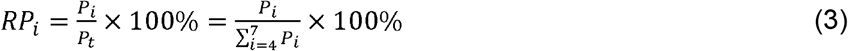

Here the *P*_*i*_ was the absolute wavelet spectral power at level *i* that equaled to the sum of squared wavelet coefficients, and *P*_*t*_ was the total absolute spectral power. The DWT relative power *RP* is an index which characterizes the distribution of brain signal powers in the four frequency bands (i.e., delta, theta, alpha, and beta) in the n-back tasks.

**Table 2.** DWT subbands information and their corresponding EEG frequency bands.

| DWT subbands | Frequency range corresponding EEG (Hz) | Corresponding frequency band (Hz) |
| --- | --- | --- |
| D5 | 15.63-30 | Beta (13-30) |
| D6 | 7.81-15.63 | Alpha (8-13) |
| D7 | 3.91-7.81 | Theta (4-8) |
| A7 | 0.5-3.91 | Delta (0.5-4) |

Among many wavelet families, the Daubechies wavelets have the properties of orthogonality and efficiency in filter implementation and were used in EEG spectral analysis(Subasi, 2007; Zarjam et al., 2015). Specifically, Daubechies wavelet with four vanishing moments (Db4) was adopted due to its suitability for analyzing EEG signals in AD in several studies (AlSharabi et al., 2022; Fiscon et al., 2018; Ghorbanian et al., 2013; Petrosian et al., 2001; Polikar et al., 2007). The Wavelet Toolbox of MATLAB (The MathWorks, Natick, Massachusetts) was employed in this study.

### 2.4 Classification approach

In order to decode group participants, we applied the multivariate pattern analysis (MVPA) (Grootswagers et al., 2017; King & Dehaene, 2014). The MVPA framework analyzes multivariates comprising multiple EEG channels or time points. MVPA usually encompasses a set of machine learning algorithms that provide an effective solution to reveal brain activation patterns associated with certain perceptual and cognitive states, specific stimuli, or other relevant information of participants.

We used the SVM machine learning method (Cortes & Vapnik, 1995) because it proved to perform well in previous AD studies compared to other classification algorithms (Chai et al., 2023; Khatri & Kwon, 2022; Menagadevi et al., 2023).

Due to the limited size of the dataset, we utilized a 5-fold cross-validation (CV) to validate the effectiveness of the classification (James, 2021). The complete datasets were randomly divided into two independent parts with an 80:20 ratio after data preparation, with 80% utilized for training and 20% used for testing. Then different data portions were used to test and train on subsequent iterations. This process was repeated until every fold of data served as the test set.

We assessed classifier performance using the receiver operating characteristic (ROC) curve analysis. The AUC (area under the ROC curve) value was used as it is more robust to imbalanced classes than the classification accuracy for binary classification (Treder, 2020). Generally, the classification performance was considered good with an AUC higher than 0.7 (Di Flumeri et al., 2018; Fawcett, 2006). The AUC metric has been adopted in several AD diagnosis studies (Callahan et al., 2015; Liu et al., 2018; Moscoso et al., 2019).

We implemented this study by employing the MVPA-Light toolbox (Treder, 2020) for the multivariate analysis of EEG signals. The wavelet-based relative power extracted from different decomposition levels (i.e., delta, theta, alpha, and beta bands) on every EEG channel were used as features to represent EEG signals. The default implementation of SVM classifiers in MVPA-Light was used. The AUC values output from the toolbox were adopted as the performance metric. The cross-validation was repeated five times with new randomly assigned folds to achieve a more stable estimate of classification. The final performance was obtained by averaging all the repetitions.

### 2.5 Statistical analysis

In order to test the difference in the wavelet-based relative power between the two groups, a two-sample unequal variance t-test was used on each channel. Four midline EEG channels from the international 10-20 system, i.e., Fz, Cz, Pz, Oz, were selected to represent the relative power distribution among the central line. A threshold probability value of p ::: .05 was used to indicate statistical significance. Multiple comparisons were not corrected due to the exploratory nature of this study.

## 3. Results

The significant differences of target condition were most apparent in the 2-back task (Table 3). Relative delta band power at the channel locations Cz (p = .05) and Pz (p = .02) was lower in the preclinical AD group compared to the control group. Conversely, relative theta (Pz, p < .01) and beta power (Fz, p = .04; Cz, p = .02; and Pz, p = .02) was higher in the preclinical group compared to the control group.

**Table 3.** Wavelet-based relative power of target condition at midline EEG channels (mean and standard deviation, unit: percent). The bold values were significant at p < .05.

|  |  | 0-back |  | 1-back |  | 2-back |  |
| --- | --- | --- | --- | --- | --- | --- | --- |
|  |  | Preclinical AD | Controls | Preclinical AD | Controls | Preclinical AD | Controls |
| delta | Fz | 67.99(15.83) | 72.43(16.85) | 70.93(17.19) | 75.27(15.13) | 72.09(12.96) | 79.31(12.70) |
|  | Cz | 67.24(15.59) | 73.50(17.47) | 71.35(15.60) | 74.03(15.44) | **71.30(13.27)** | **79.93(14.47)** |
|  | Pz | 79.71(13.37) | 82.78(9.38) | 79.95(11.14) | 84.06(11.92) | **73.61(17.25)** | **85.06(11.25)** |
|  | Oz | 79.87(11.79) | 79.64(8.15) | 77.15(13.89) | 82.93(10.20) | 76.83(13.42) | 78.90(12.57) |
| theta | Fz | 16.87(10.80) | 14.78(11.79) | 17.09(13.12) | 14.82(10.59) | 16.66(10.87) | 11.76(7.56) |
|  | Cz | 17.32(9.79) | 13.72(11.53) | 17.56(14.42) | 13.60(10.38) | 16.73(11.24) | 11.27(9.86) |
|  | Pz | **9.69(5.25)** | **6.54(3.99)** | 9.81(7.36) | 6.16(6.61) | **11.48(8.45)** | **5.05(3.88)** |
|  | Oz | 8.83(5.92) | 9.74(7.02) | 10.09(7.69) | 6.53(4.01) | 9.51(7.02) | 8.20(5.99) |
| alpha | Fz | 11.08(7.46) | 9.84(7.42) | 8.68(8.45) | 7.50(6.03) | 7.09(4.23) | 6.36(6.80) |
|  | Cz | 11.00(8.81) | 10.21(7.40) | 7.98(6.25) | 9.66(6.96) | 8.11(4.89) | 6.43(5.59) |
|  | Pz | 8.18(9.07) | 7.78(4.77) | 6.82(3.63) | 7.41(5.85) | 9.68(8.63) | 7.33(7.88) |
|  | Oz | 8.61(7.19) | 7.60(3.93) | 8.88(6.30) | 7.43(5.88) | 8.93(6.27) | 9.25(9.24) |
| beta | Fz | 4.07(3.57) | 2.96(2.06) | 3.30(1.76) | 2.42(2.04) | **4.16(2.70)** | **2.57(2.05)** |
|  | Cz | **4.45(2.71)** | **2.58(1.63)** | 3.11(1.93) | 2.71(2.46) | **3.86(2.21)** | **2.37(1.55)** |
|  | Pz | 2.42(1.67) | 2.89(1.89) | 3.42(2.48) | 2.37(2.06) | **5.23(4.82)** | **2.55(1.60)** |
|  | Oz | 2.69(2.37) | 3.02(1.16) | 3.88(2.84) | 3.12(2.45) | 4.72(4.01) | 3.65(2.26) |

Fewer significant differences were observed in the 0-back and 1-back tasks. In the 0-back task, participants with preclinical AD showed greater power compared to controls in the theta band at channel Pz (p = .04) and in the beta band at channel Cz (p = .01).

The relative power of the nontarget condition in Table 4 showed a similar pattern to the target condition in Table 3.

**Table 4.**
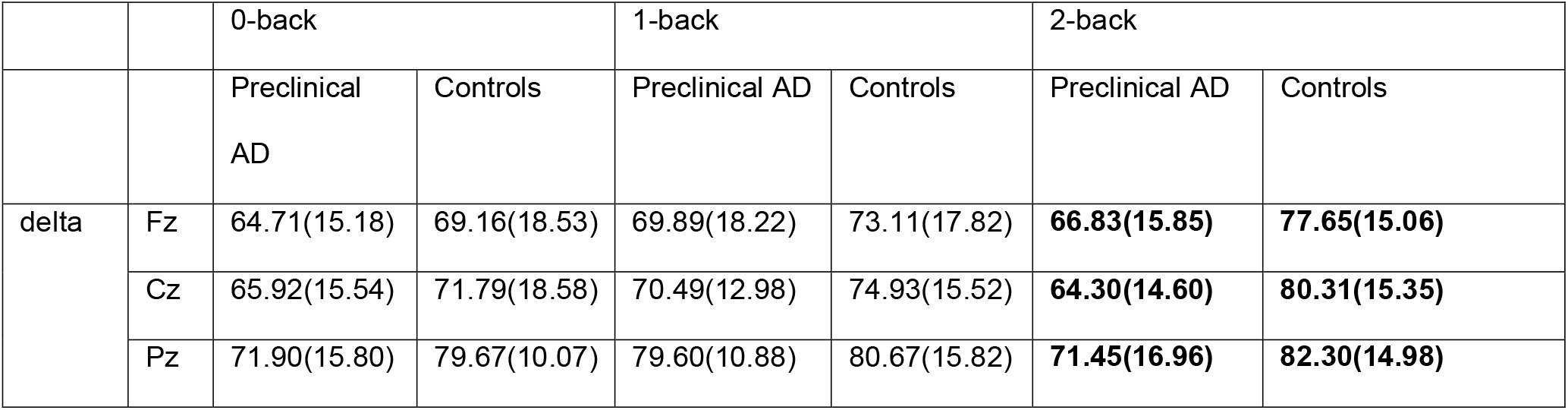
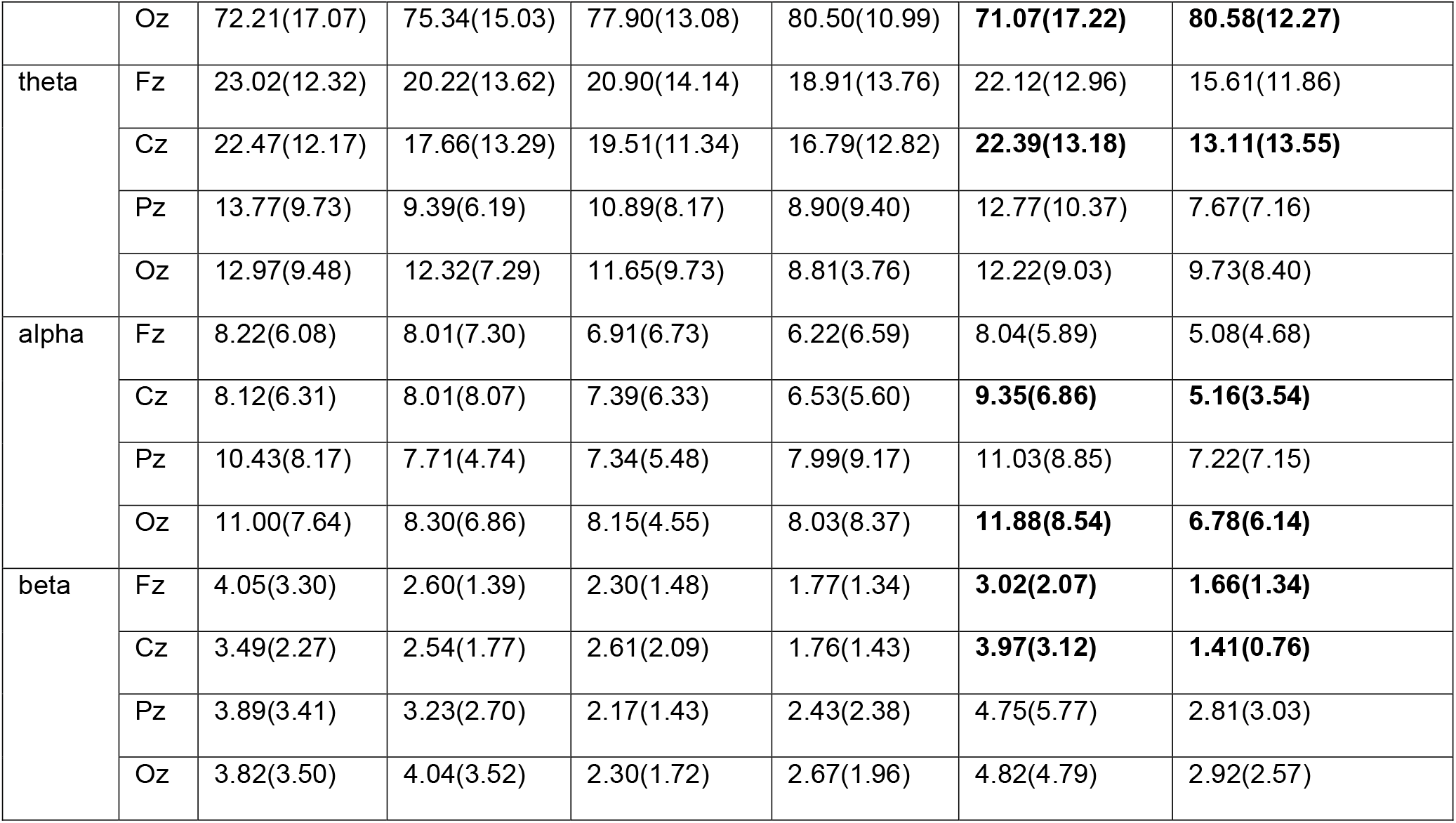
Wavelet-based relative power of nontarget condition at midline EEG channels (mean and standard deviation, unit: percent). The bold values were significant at p < .05.

Relative power in the delta band was lower in the preclinical group compared to the control group, while the reverse was true in theta, alpha, and beta bands at the midline channels. In the 2-back task, the preclinical group exhibited lower delta band power at all channels: Fz (p = .03), Cz (p < .01), Pz (p = .04), and Oz (p = .05). The power was significantly higher in the preclinical group than the controls in the theta band at Cz (p= .03), in the alpha band at Cz (p = .02) and Oz (p = .03), and in the beta band at Fz (p = .02) and Cz (p < .01).

None of the channels showed a significant difference between the two groups in the 0-back and 1-back tasks.

The AUC topographies showed higher values in the 2-back compared to the 0-back and 1-back tasks (Figure 2). The 0-back and 1-back tasks had similar AUC performances, with no consistent AUC contours in these tasks.

**Figure 2.**
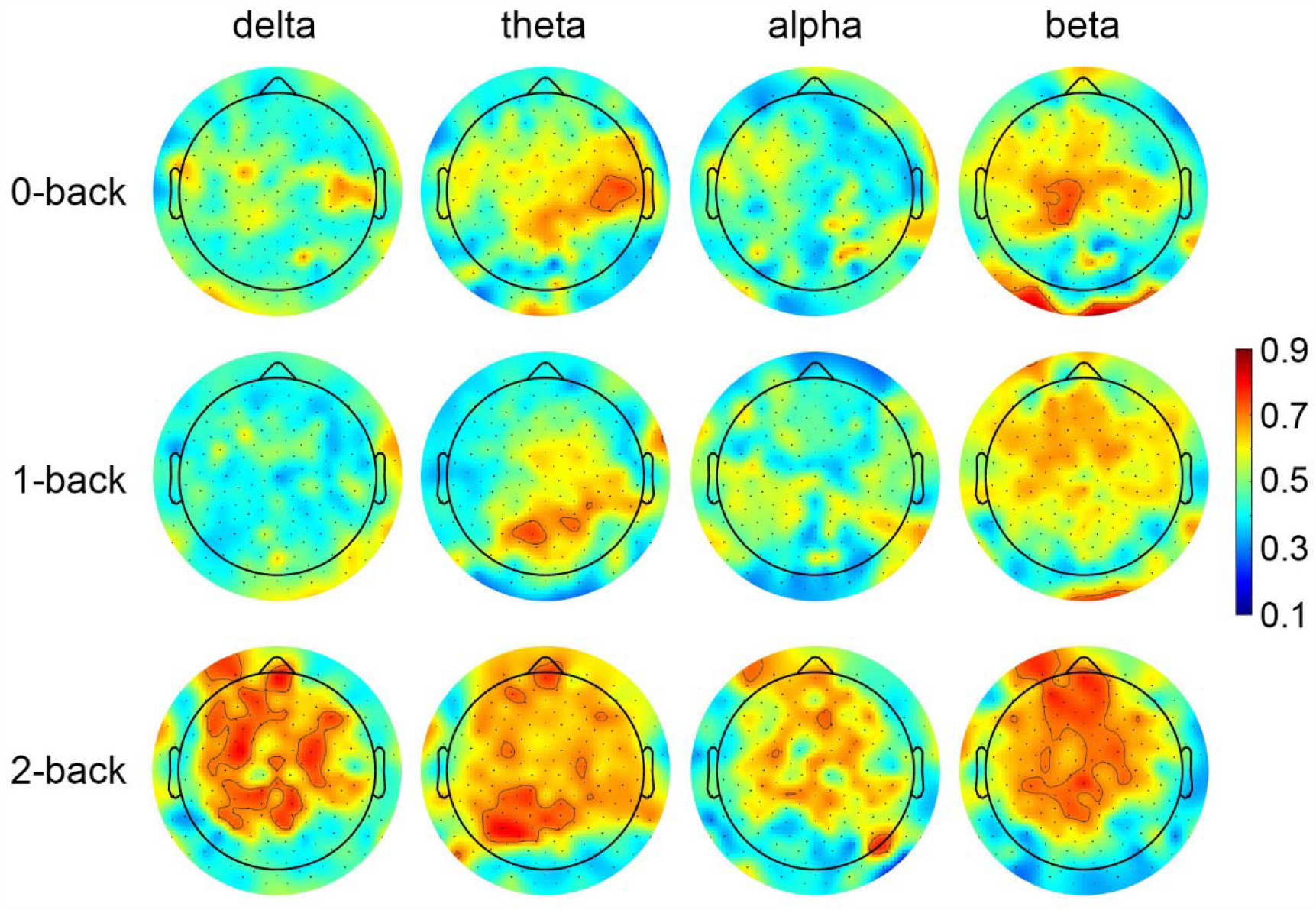
Topographies of AUC values of target condition in the 0-back, 1-back, and 2-back tasks. Channels with AUC higher than 0.7 were contoured.

Figure 2 shows AUC value topographies of all frequency bands (i.e., delta, theta, alpha, and beta) of the n-back target condition.

In the 2-back task, high AUC values (>0.7) were observed in the frontal, central, and parietal areas. Specifically, Table 5 showed that the highest AUC values in the 2-back task were observed in the theta (AUC = 0.81) and delta bands (AUC = 0.80), followed by the beta band (AUC = 0.77). The alpha band was the lowest (highest AUC = 0.72).

**Table 5.**
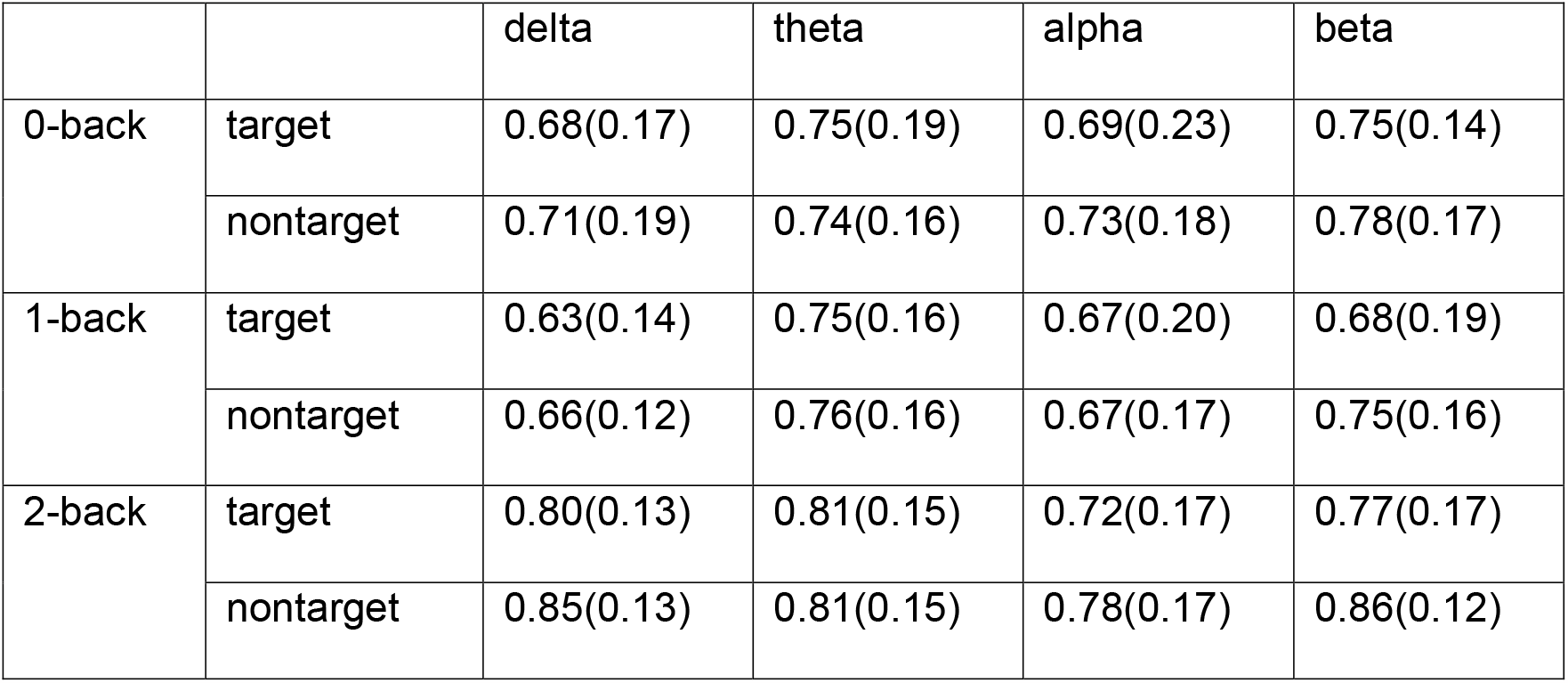
The highest AUC values of all frequency bands in n-back tasks for target and nontarget conditions (mean with standard deviation).

|  |  | delta | theta | alpha | beta |
| --- | --- | --- | --- | --- | --- |
| 0-back | target | 0.68(0.17) | 0.75(0.19) | 0.69(0.23) | 0.75(0.14) |
|  | nontarget | 0.71(0.19) | 0.74(0.16) | 0.73(0.18) | 0.78(0.17) |
| 1-back | target | 0.63(0.14) | 0.75(0.16) | 0.67(0.20) | 0.68(0.19) |
|  | nontarget | 0.66(0.12) | 0.76(0.16) | 0.67(0.17) | 0.75(0.16) |
| 2-back | target | 0.80(0.13) | 0.81(0.15) | 0.72(0.17) | 0.77(0.17) |
|  | nontarget | 0.85(0.13) | 0.81(0.15) | 0.78(0.17) | 0.86(0.12) |

The AUC topographies of the nontarget condition, shown in Figure 3, followed a consistent pattern as in Figure 2, with the 2-back task achieving higher AUC values than the 0-back and 1-back tasks. The delta and beta bands had the highest AUCs (0.85 in the delta band and 0.86 in the beta band, 2-back data, Table 5), followed by the theta band (AUC = 0.81). The alpha band had the lowest value (highest AUC = 0.78). The contours of channels with AUC higher than 0.7 covered the frontal, central, and parietal brain areas (2-back task).

**Figure 3.**
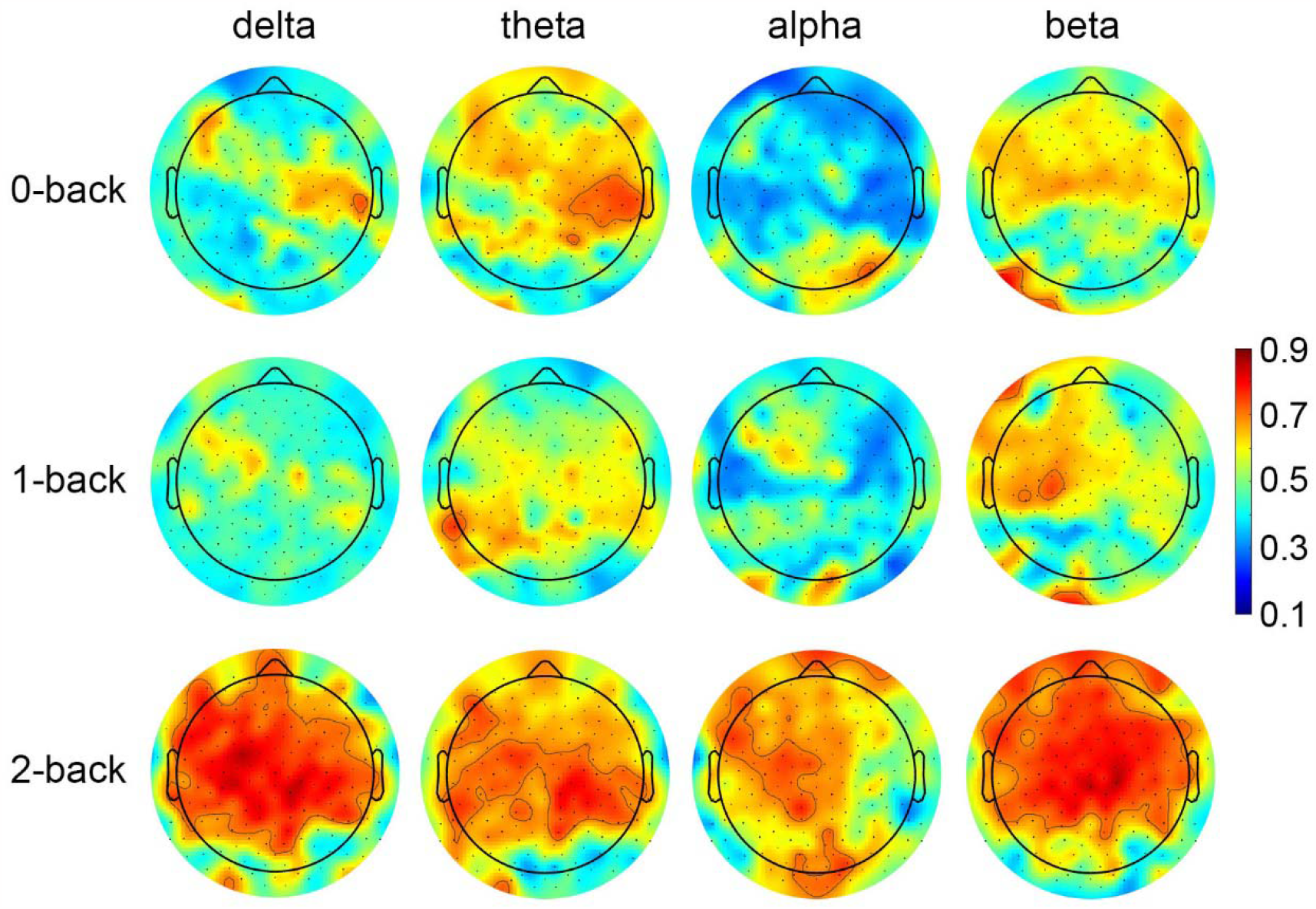
Topographies of AUC values of nontarget condition in the 0-back, 1-back, and 2-back tasks. Channels with AUC higher than 0.7 were contoured.

Table 5 shows the highest AUC values of all frequency bands in 0, 1 and 2-back tasks.

The AUC of the 2-back task grossly exhibited higher values than the 0-back and 1-back tasks in each frequency band for both target and nontarget conditions.

The nontarget condition generally obtained higher AUC values than the target condition. The highest AUC values were from 2-back nontarget beta band data (AUC = 0.86), followed by the delta band (AUC = 0.85).

## 4. Discussion

The outcomes of this study underscore the utilization of machine learning approaches, employing EEG spectral features from task-based data, to distinguish cognitively normal older adults with preclinical AD from cognitively normal controls. While a few studies have noted task-related EEG modifications in older adults with an elevated risk of AD (Arakaki et al., 2019; Choi et al., 2023), these investigations have not incorporated machine learning techniques for group classification. Our study addresses this gap by providing validation that memory-related EEG oscillations can signify neurocognitive changes preceding clinical diagnosis. Furthermore, we establish that these EEG patterns encompass substantial information for the early identification of older adults at risk of AD.

The highest classification performance of this study was obtained in the beta band (AUC = 0.86) and the delta band (AUC = 0.85), which were almost identical (2-back task, nontarget data in Table 5). Both beta and delta oscillations are believed to be related to working memory function (Harmony, 2013; Harmony et al., 1996; Miller et al., 2018; Schmidt et al., 2019; Spitzer & Haegens, 2017). Our results revealed that the beta and delta EEG bands carried the most information on spectral changes between adults with preclinical AD and controls. This is in line with Fernández et al. (2006), who identified that relative power in the beta (16-28 Hz) and the delta (2-4 Hz) frequency bands were the most sensitive variables in differentiating AD patients and controls.

Our finding that the delta-band relative power of the 2-back task in the preclinical AD group was lower than the control group was in accordance with our previous study (Devos et al., 2022), as well as the relative power increase of other frequency bands in the preclinical AD group at Cz, Pz, and Oz channels (Tables 3 and 4). The overall spectral shift towards decreased power in the low-frequency delta band and increased power in the high-frequency beta bands in the preclinical AD group are consistent with neural hyperexcitability, a finding increasingly reported in the literature (Devos et al., 2022; Targa Dias Anastacio et al., 2022). Although the drivers of this increased neural excitability in this preclinical phase of AD are still unclear (Targa Dias Anastacio et al., 2022), studies in animal models of AD have found a strong link between hyperexcitability and Aβ accumulation near the hyperexcitable neurons (Busche & Konnerth, 2016). Previous EEG studies also reported that individuals with MCI have higher power from theta to beta bands than controls during working memory tasks (Jiang, 2005). This finding aligns with our results on theta, alpha, and beta bands power, in which the preclinical AD group shows larger values than the control group, and most of the significant differences may be elicited with more difficult working memory tasks such as the 2-back task (Tables 3 and 4).

The AUC topographies showed higher values in the front-centro-parietal brain areas (Figures 2 and 3, delta and beta bands, 2-back task). This region included the frontal lobe, where the event-related potential (P3 component) was found to provide good reliability for studying the effect of Aβ on neural transmission in preclinical AD in our previous study (Devos et al., 2020) since the frontal lobe is tightly associated with working memory (Kondo et al., 2004). One study that identified amnestic MCI using convolutional neural networks also showed that the channels in the frontal lobe were with a top representation of patients’ cognitive states (Li et al., 2022). The possible mechanism of delivering high AUCs in the front-centro-parietal areas is that working memory relies on different functional sub-systems, and it is attributable to the coordination of disparate brain areas, in particular frontal and parietal regions (Imaruoka et al., 2005; Owen et al., 2005).

When comparing performance between tasks, the 2-back task achieved higher AUCs than the 0-back and 1-back tasks (Table 5). Previous studies suggested that different work or memory load levels could invoke spectral power alterations in various frequency bands (Baldwin & Penaranda, 2012; Brouwer et al., 2012; Christensen et al., 2012; Michels et al., 2010; Pesonen et al., 2007). We assume that the high workload task, i.e., 2-back, might have the potential to fully exploit the working memory capacity and produce more discriminatory EEG oscillatory activity patterns between the preclinical AD and control groups.

In the current study, the AUC of the classifier with the best performance was 0.86. Previous studies reported highest AUC values ranging between from 0.80 to 0.89 in classifying AD vs. controls (Babiloni et al., 2016; Chai et al., 2019; Chedid et al., 2022; de Haan et al., 2008; Ding et al., 2022; Fernández et al., 2006; Meghdadi et al., 2021; Trambaiolli et al., 2011), and between 0.60 and 0.81 in classifying MCI vs. controls (Ding et al., 2022; Gómez et al., 2009; Meghdadi et al., 2021; Sibilano et al., 2023). Considering that those studies investigated disease stages of AD with apparent cognitive impairments, our model achieved similar classification performance in a preclinical phase of AD with normal cognition. Our results suggest that electrophysiological changes could occur prior to cognitive changes, and the event-related oscillations could serve as easy, relatively inexpensive, yet accurate markers of preclinical AD.

Although our study compares well with the participant size of previous studies (Chai et al., 2019; Gómez et al., 2009; Mazaheri et al., 2018; Trambaiolli et al., 2011), we acknowledge the limitation that the size of the participants is small, and a future study with larger-scale samples is needed. Furthermore, EEG power spectra could change during the course of disease. In a 2.5-year follow-up study of AD, an increase of delta and theta activities and a decrease of alpha and beta activities were reported (Coben et al., 1985). A longitudinal study is warranted to confirm the use of EEG as an accurate marker of cognitive decline. Finally, the time interval between PET and EEG scans was relatively long. We cannot rule out that some participants with preclinical AD might have converted to MCI. Nevertheless, the average MOCA scores were higher than 26, indicating no cognitive impairments in the preclinical AD group.

## 5. Conclusion

In this study, an SVM-based machine learning model was built to discriminate preclinical AD and neurotypical older adults, using EEG data from n-back working memory tasks. The event-related oscillation was analyzed in all frequency bands using features extracted from discrete wavelet transform. Our results showed that relative power from the beta and delta bands, particularly in the 2-back task, can achieve higher classification performance than other frequency bands and less demanding tasks. This study demonstrates that machine learning based on memory-related power spectral features are capable of characterizing the brain activities change associated with preclinical AD from healthy controls. Our results imply that EEG oscillations may reveal pathophysiological changes in the earliest stage of AD when no cognitive impairments are apparent.

## Data Availability

All data produced in the present study are available upon reasonable request to the authors

## Notes

### Competing Interest Statement

The authors have declared no competing interest.

### Funding Statement

This work was completed at the Hoglund Biomedical Imaging Center which is supported by the Forrest and Sally Hoglund and a High-End Instrumentation grant from the National Institutes of Health (S10 RR29577). Research reported in this publication was supported by the National Institute on Aging of the National Institutes of Health under Award Number K01 AG058785 and P30 AG072973. This study was supported in part by a pilot grant of the KU Alzheimer Disease Research Center (P30 AG035982). The content is solely the responsibility of the authors and does not necessarily represent the official views of the National Institutes of Health.

### Author Declarations

IRB of University of Kansas Medical Center gave ethical approval for this work.

